# Covid-19 Outbreak Progression in Italian Regions: Approaching the Peak by March 29^th^

**DOI:** 10.1101/2020.03.30.20043612

**Authors:** Cosimo Distante, Prisco Piscitelli, Alessandro Miani

**Affiliations:** CNR ISASI, Institute of Applied Sciences and Intelligence Systems, Lecce, Italy; Euro Mediterranean Scientific Biomedical Institute (ISBEM), Bruxelles, Belgium; Italian Society of Environmental Medicine (SIMA), Milan, Italy; Department of Environmental Science and Policy, University of Milan, Milan, Italy

## Abstract

**Background:** Italy and especially the Lombardy region is experiencing a heavy burden of Covid-19 infection. The peak of the epidemics has not yet been reached and it is expected to be delayed in Central and Southern Italian regions compared to Northern ones. We have modeled the Covid-19 outbreak progression in Italian Regions vs. Lombardy.

**Methods:** In our models, we have estimated the basic reproduction number (R_0_) -which represents the average number of people that can be infected by a person who has already acquired the infection - both by fitting the exponential growth rate of the infection across a 1-month period and also by using day by day assessment, based on single observations. We used the susceptible–exposed–infected–removed (SEIR) compartment model to predict the spreading of the pandemic in Italy.

**Results:** The two methods provide agreements of values, although the first method based on exponential fit should provide a better estimation, being computed on the entire time series. Taking into account the growth rate of the infection across a 1-month period, in Lombardy each infected person has involved other 5 people (4.94 base on data of March 22^nd^ vs. 5.07 based on data of March 19^th^) compared to a value of *R*_0_ = 2.68 reported in the Chinese city of Whuan. According to our model and Piedmont, Veneto, Emilia Romagna, Tuscany and Marche reach an R_0_ value up to 4. The R_0_ is 3.7 for Lazio and 3.6 for Campania region, where this latter shows the highest value among the Southern Italian regions, followed by Apulia (3.5), Sicily (3.4), Abruzzo (3.4), Calabria (3.1), Basilicata (2.5) and Molise (2.4). The value of R_0_ is decreasing in Lombardy and Northern Regions, while it is increasing in Central and Southern Regions.

**Conclusion:** The expected peak of SEIR model can be forecast by the last week of March at national level, and by the first weeks of April in Southern Italian Regions. These kind of models can be useful for adoption of all the possible preventive measures, and to assess the epidemics progression across Southern regions as opposed to the Northern ones.

## Epidemiological figures

According to the Italian National Institute of Health (ISS), at the date of March 19^TH^ in Italy there were 35731 people who were tested positive to Wuhan novel coronavirus (2019-nCoV). Men were 20686 and women were 14378. Women are the majority of cases in the age group 20-29 (n=732 females vs. 603 males) and >90 years old (n=694 females vs. 397 males); women and men are equally distributed between 30-39 and 40-49 years old (n=1202 females vs. 1261 males, and n=2123 females vs.2186 males, respectively), while men represent the majority of casesin all the other age groups (n=118 vs 84 from 0 to the age of 9; n=150 vs. 117 between 10 and 19 years old; n=3775 vs. 2926 from 50 to 59 years of age; n=4221 vs. 2025 from 60 to 69; n=4702 vs. 2290 from 70 to 79; n=3158 vs. 2100 from 80 to 89).[7] Regional figures are available up to March 23rd and show that about 18910 out of 50418 currently positive people were detected only in Lombardy (the region of Milan), followed by Emilia Romagna (n=7220), Veneto (n=4986), Piedmont (n=4529), Marche (n=2358), Tuscany (n=2301), Trentino Alto Adige (n=1602), Liguria (1553), Lazio (n=1414), Campania (n=929), Apulia (n=862), Friuli Venezia Giulia (n=771), Sicily (n=681), Abruzzo (n=605), Umbria (n=556), Val d’Aosta (n=379), Sardinia (n=343), Calabria (n=280), Basilicata (n=89) and Molise (n=50). [7]

A total of 20692 symptomatic people were hospitalized at the same date of March 23^st^ in Italy, with Lombardy accounting for 9266 hospital admissions, followed by Emilia Romagna (n=2846), Piedmont (n=2194) and Veneto (n=1206). Only Tuscany (n=838), Marche (n=882), Liguria (n=761) and Lazio (n=718) recorded more than 250 hospital admissions at regional level, while other regions remain still lower. At the present, 3204 patients are assisted in intensive care units (1183 in Lombardy, 343 in Piedmont, 281 in Veneto, 276 in Emilia Romagna, 238 in Tuscany, 148 in Marche region and 133 in Liguria. Except Campania (n=110) and Lazio (n=96), all the other regions of Central and Southern Italy, at the moment have less than 50 patients admitted to the ICUs of their regional healthcare systems. [7]

On March 23^rd^, total deaths were 6077 at national level (more than the overall 3264 deaths observed in China), with 3776 in Lombardy, 802 in Emilia Romagna, 315 in Piedmont, 212 in Liguria, 203 in Marche region, 192 in Veneto, 109 in Tuscany, 70 in Trentino Alto Adige, 63 in Lazio, 54 in Friuli Venezia Giulia, 49 in Campania, 38 in Abruzzo, 37 in Apulia, and less than 15 in the other six regions (Figure 1).[7]Lethality rates seems to increase with age and it is higher in males: 0% from 0 to 29 and 0,5% between 30 and 49 years of age; 1.2% in the age group 50-59 (0.6% in women and 1.7% in men); 4.9% from 60 to 69 years old (2.8% in women and 6.0% in men); 15.3% from 70 to 79 (10.7% in females and 17.8% in males); 23.2% from 80 to 89 years old (19.1% and 26.4% in men).[7]

**Figure 1.**
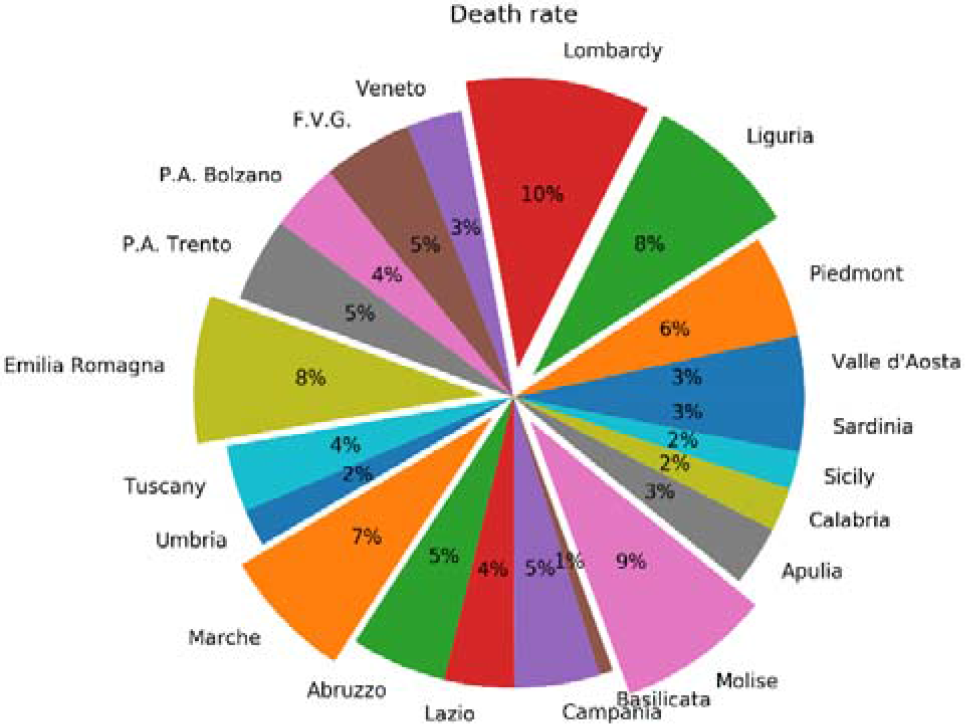
Death rate in Italy per region at the date of March 23^rd^.

Based on these figures, it is clear that the Covid-19 outbreak is now putting overwhelming pressure mainly on Lombardy and Northern regions of the Po Valley (Padana plain), but the peak of the epidemics has not yet been reached. Until now, Southern Regions seem to be less affected by the Covid-19 infection although a huge number of people - mainly students attending Universities in Northern Italy - came back from Po Valley to their families in the South just in the middle of the outbreak, thus representing a potential factor able to accelerate the spreading of the viral infection. Here we present an attempt to predict the peak of the outbreak in Italy, which is expected at national level by the end of March, and the different progression of the epidemics in Southern Italian Regions compared to Lombardy.

## Modeling the Covid-19 outbreak progression in Southern Italian Regions vs. Lombardy

The basic reproduction number (R_0_) is an indicator that resumes the average number of people that can be infected by a person who has already acquired the infection. R_0_ is a metric of how contagious is the disease and its correct estimation is extremely important for epidemiologists, especially when facing new diseases like COVID-19. R_0_ can be computed in different ways. In our models, we have estimated the basic reproduction number (R_0_) both by fitting the exponential growth rate of the infection across a 1-month periodand also by using day by day assessment, based on single observations [1]. This study makes use of the susceptible–exposed–infected–removed (SEIR) compartment model [4] to predict the spreading of the pandemic in Italy. Our efforts could be helpfulin the adoption of all the possible preventive measures, and to study of the epidemics progression across Southern regions as opposed to the national trend. This metric can be biased by the optimal estimation of the basic reproductive number R_0_ (pronounced R-nought). It must be said that R_0_ is important if correlated with weather conditions and that reproductive index is reduced as the air temperature and relative humidity increase,[5] according to the formula:

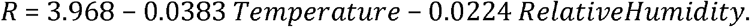

This means the transmission of COVID-19 candecrease with the warmer season, and that at least some specific figures of the outbreak in Lombardy and Po Valley can be explained taking into account climatic variables.

## Modeling basic reproductive number R_0_

### Exponential framework estimation

The exponential estimation is based on the work of Wu et al[7], which essentially was based on that of Zhao and colleagues[8], where the epidemic curve obey to the exponential growth.

At the date of this study (March 23^rd^2020), the epidemic growth was still near exponential growth, and the fitted model has many inliers data points. The method is based on non-linear least square framework for intrinsic growth estimation, in order to obtain with M the Laplace transforming the probability distribution of the serial interval of the infection. The estimation is obtained with 100% susceptibility for 2019-nCoV at the early stage in Wuhan as reported in [7]. In Figure 2, the number estimates are computed for the Southern Italian regions and for the initialoutbreak region (Lombardy). According to our model, in Lombardy, each infected person has involvedother 4.9 people (5.07). The R_0_ lowers to 3.6 for Campania region, which shows the highest value among the Southern Italian regions, followed by Apulia (3.5), Sicily (3.4), Abruzzo (3.4), Calabria (3.2), Molise (2.4), and Basilicata (2.2).

**Figure 2.**
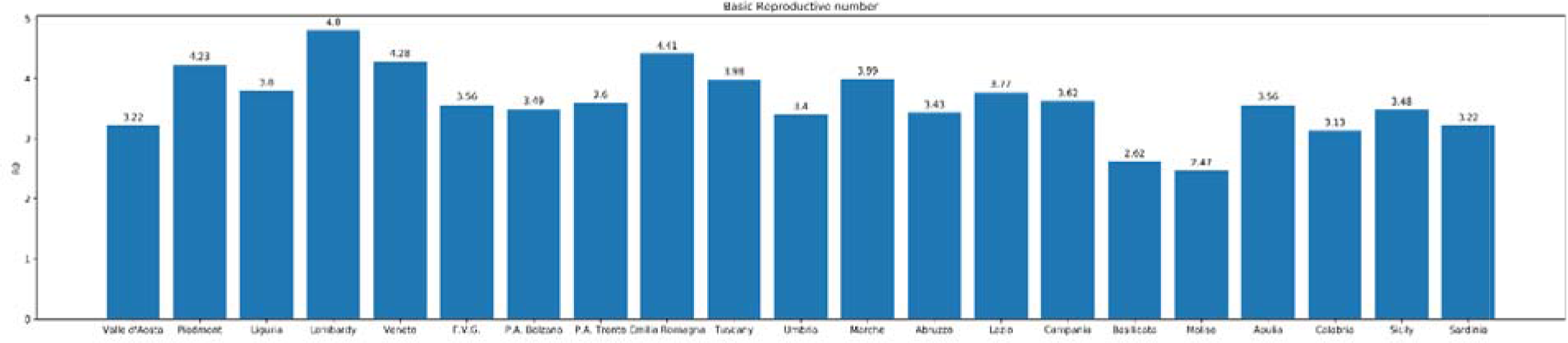
Basic Reproductive index computed with exponential fitting method along the entire time series (1-month period).

### Daily basis estimation of the reproductive number

R_0_ is an average value, but it can be also computed day by day to monitor the transmission of the infection. Being an average value, it can be skewed by super-spreader events. A super-spreaderis an infected individual who infects an unexpectedly large number of people. In Italy this event can be also generated not necessarily by an individual, but from the perturbation of a susceptible population, as it happened in Apulia and Sicily with uncontrolled large group of people coming from outbreak areas. For a “super spreader” individual, such events are not necessarily a bad sign, because they can indicate that fewer people are perpetuating an epidemic. Super-spreaders may also be easier to identify and contain, since their symptoms are likely to be more severe. In short, R_0_ is a moving target. Tracking every case and the transmission of a disease is extremely difficult, so the estimation of R_0_ is a complex and challenging issue: estimates often change as new data becomes available. If we define the Y(t) as the number of infected people with symptom at time t, the exponential growth rate is. Let us consider the generation time (i.e. the serial interval) and the latent or incubation time (values taken from [6]). The infectious time, and the ratio of exposed period to generation time is. The basic reproductive number can be approximated to:

In order to estimate, it is important to find and then the number of infected people:

Suspects correspond to the number of individuals screened with test which have been confirmed. Figure 3 and Table 1 show the estimated values, computed on a daily basis for the Italian regions and for the initial outbreak region Lombardy, where about 40 cases were confirmed out of 100 suspects (Figure 4).The two methods provide agreements of values, although the first method based on exponential fit should provide a better estimation, being computed on the entire time series. From Figure 3, it comes an important aspect with respect to the Wuhan as reported in [6].

**Table 1.**
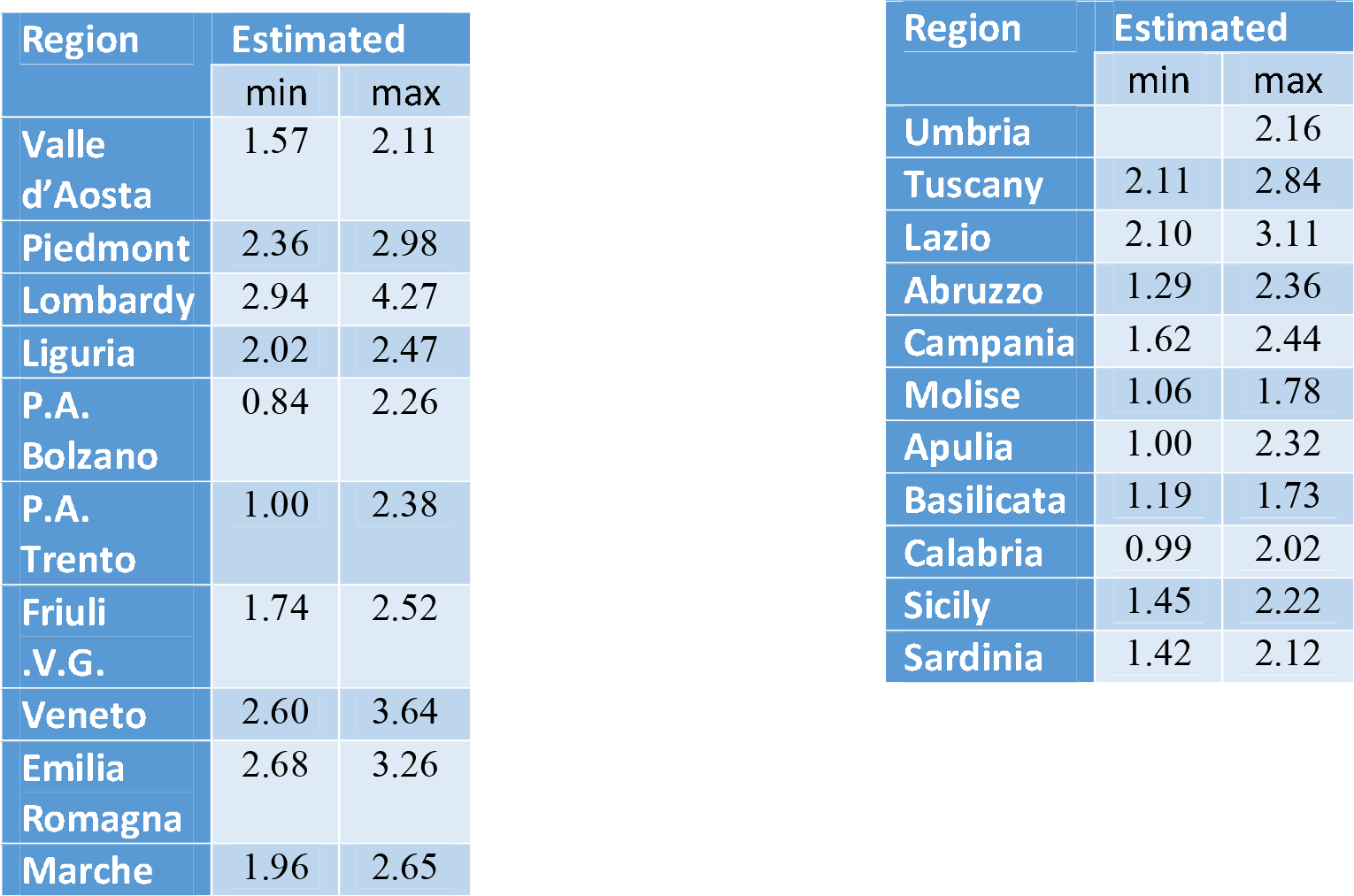
values of the analyzed regions. Given the trend, min values are usually referred to the beginning of the infection

**Figure 3.**
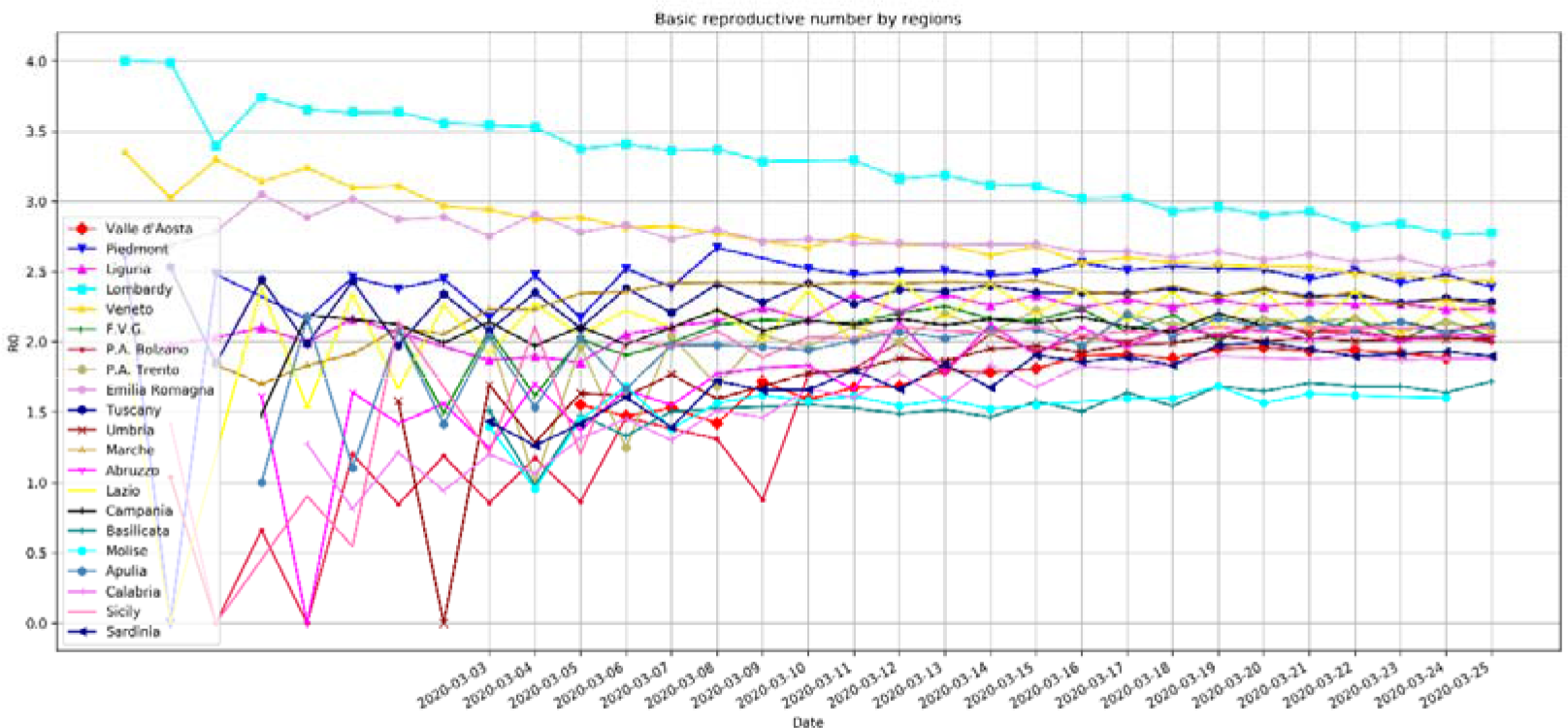
Basic reproductive index computed on a daily basis with the second method investigated in this letter.

**Figure 4.**
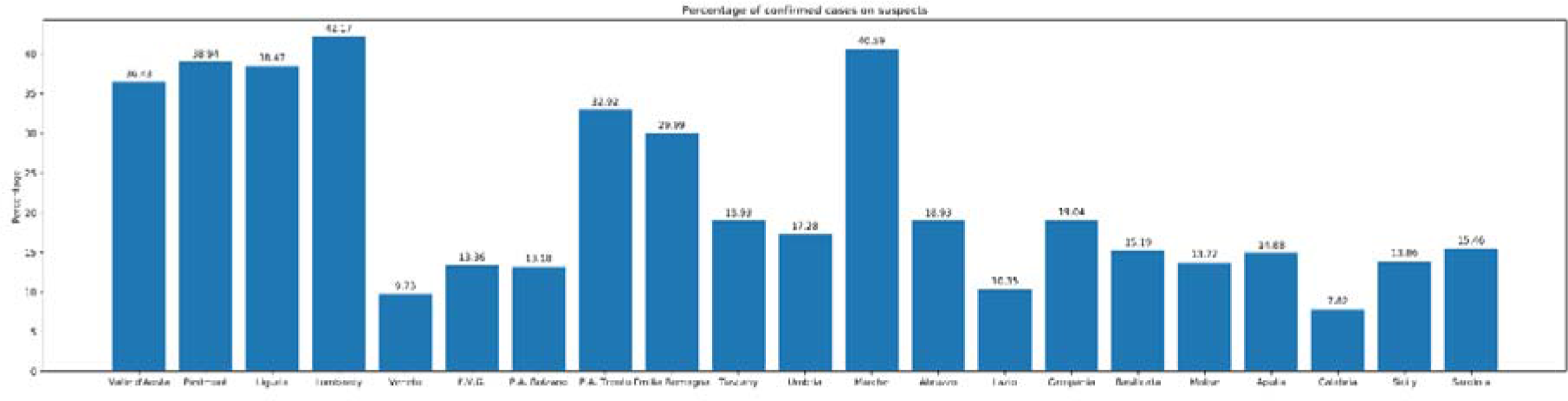
Percentage of suspects confirmed at the time of March 23^rd^ hours. In one day, Lombardy increased by 10% with respect to the previous 24 hours.

### Modeling transmission in Italy

We used the susceptible-exposed-infectious-recovered (SEIR) model [4] to simulate epidemic since it was established on January 2020. It is based on a previous model SIR which was based on three compartments, but since the infection has an incubation period, the compartment E (Exposed) is included. This compartments are modeled over the time, and capture the changes in the population. Let us say that given N the total population, then N=S+E+I+R, where:

- “S” Susceptible is the portion of population that does not have any vax coverage or immune;
- “E” exposed: is the portion of the population that have been infected, but are in the incubation period that do not infect others;
- “I” Infectious: is the portion of N that is infectious and may infect others, they become dead or may recover;
- “R” Recovered: number of infectious people who have been healed, and become immune.

**Figure 2.**
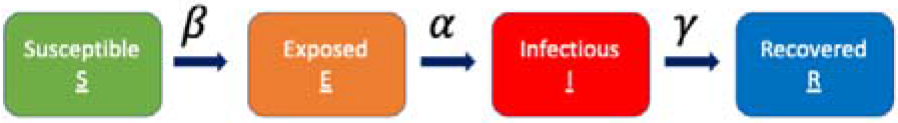
SEIR model with the four compartments and their relationships

This model captures dynamics of these compartments over the time by four ordinary differential equations. One of the most important aspects of these ordinary differential equations is equilibrium, who is achieved by setting to zero their derivatives along the time t. The two equilibriums are: disease-free equilibrium (DFE) and endemic equilibrium (EE).

Besides equilibrium, stability is an issue who is correlated with the basic reproductive number, where if *R*_0_ <1 DFE is stable; while when *R*_0_ >1 DFE is unstable and EE is stable. The four equations are:

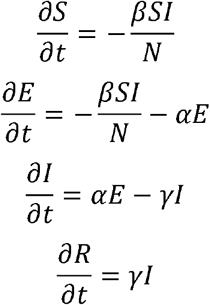

Where *β* = *R*_0_/*T*_*i*_, *α* = 1/*T*_*l*_ and *γ* = 1/*T*_*i*_ with *T*_*i*_ and *T*_*l*_ as defined above, the serial and incubation period respectively. The contact rate *β* is the rate of infection from an infected individual to one of their susceptible contacts on the unitary time step *dt*. The number of individuals transferred from Susceptible state to Exposed state is 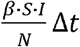. The force of infection is defined as *β * S(t)/N* which is the number of new infections divided the Italian population. At the same time step, there are *αE(t)Δt* number of cases that are transferred from Exposed to Infectious compartment, and *γ(t)Δt* number of cases transferred from Infectious compartment to Removed. It is important to state that we assume a closed population, which means the population is fixed, then no births, no deaths or introduction of new individuals. From the above ODE system, 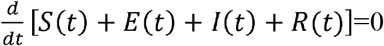 which means that the population N is constant at any time step *t: S(t) + E(t) + I(t) + R(t) = N* for any *t* ≥ 0.The individuals in exposed state is is infected but not yet infectious. The population is well mixed, and the model assumes that latent and infectious times of the pathogen are exponentially distributed. In this letter, contact rate *β* is changing over time as it happens in COVID-19, which increase in early stage due to public unawareness of the disease, then decrease with government control policy measures. The contact rate follow a logistic function trend by estimating it day by day [9]:

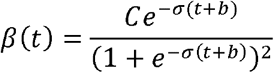

With *t* the number of days after January 31^st^ (the first found cases in Italy), *σ* a regularization parameter, and *b* the bias. A training procedure is performed on the observable data in order to find optimal *(C, σ b)*. We have considered as Exposed people, a number of twice Infected people, after lockdown which is in line with the predictions and the observed values. As shown in Figure 6 (red curve), the expected peak of SEIR model is around March 29^th^ at national level. It is expected that Southern Italian Regions could reach the peak later, in the first days of April.

**Figure 3.**
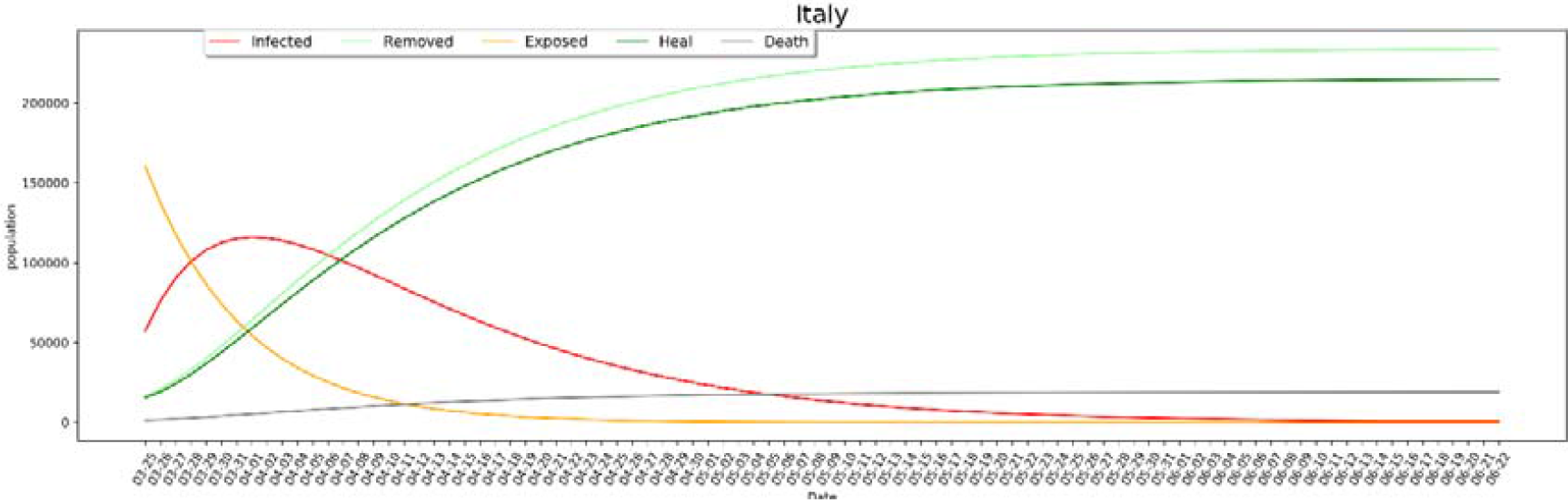
Prediction of the Exposed, Infected and death people. Peak of more than 58K infected people is expected around March 25^th^.

## Data Availability

DATA WILL BE MADE AVAILABLE UPON REASONABLE REQUEST

